# Structural brain characteristics of current co-occurring chronic pain and depression: a cross-sectional analysis of UK Biobank

**DOI:** 10.64898/2026.04.02.26350033

**Authors:** Hannah Casey, Mark J. Adams, Andrew M. McIntosh, Marie T. Fallon, Daniel J. Smith, Rona J. Strawbridge, Heather C. Whalley

**Author notes:** Correspondence to: Hannah Casey, Email address, Address: FU214, Chancellor’s Building, The University of Edinburgh, 49 Little France Cres, Edinburgh, EH16 4SB, UK.

## Abstract

**Background:** Chronic pain and depression are prevalent and burdensome conditions that frequently co-occur. Separate neuroimaging studies of each disorder suggest overlapping brain-structure alterations, however, relatively few studies have examined their comorbidity directly, and the neuroanatomical profile of co-occurring chronic pain and depression remains unclear.

**Methods:** Using UK Biobank data (n = 71,214), we conducted cross-sectional pairwise association analyses of brain structure (cortical measures, subcortical volumes, and white matter microstructure) comparing participants with current comorbid chronic pain and depression, current chronic pain only, current depression only, and controls.

**Results:** Compared with controls, the comorbidity group showed regional differences in cortical surface area and thickness (β range = −0.096 to 0.098, p_FDR_ < 0.05), widespread lower cortical volume (β range = −0.096 to −0.050, p_FDR_ < 0.05), lower thalamic (left: β = −0.048, p_FDR_ = 0.038; right: β = −0.060, p_FDR_ = 0.007), hippocampal (left: β = −0.062, p_FDR_ = 0.035; right: β = −0.088, p_FDR_ = 0.002) and left accumbens volume (β = −0.073, p_FDR_ = 0.011), and evidence of widespread white matter microstructure alterations (fractional anisotropy: β range = −0.116 to −0.080, p_FDR_ < 0.05; mean diffusivity: β range = 0.063 to 0.137, p_FDR_ < 0.05). Pairwise comparisons with the disorder-specific groups also identified several alterations unique to the comorbidity group. Compared to controls, those with chronic pain only had widespread lower cortical surface area and volume (β range = −0.043 to −0.015, pFDR < 0.05), whereas non-comorbid depression showed more regionally specific lower cortical thickness and volume (β range = −0.140 to −0.062, pFDR < 0.05) and lower thalamic volume (left: β = −0.067, p_FDR_ = 0.016; right: β = −0.066, p_FDR_ = 0.015), alongside widespread white matter microstructure deficits (fractional anisotropy: β range = −0.104 to −0.083, p_FDR_ < 0.05; mean diffusivity: β range = 0.079 to 0.149, p_FDR_ < 0.05).

**Conclusion:** These results provide a robust characterisation of brain structure alterations in comorbid chronic pain and depression, highlighting a distinct neuroanatomical profile and advancing understanding of underlying neurobiology.

## Introduction

Chronic pain and depression are common comorbidities, with 39.3% of adults with chronic pain estimated to have clinically significant depressive symptoms (Aaron et al., 2025). Both disorders are large global health burdens and represent leading causes of disability worldwide (Hay et al., 2025). Their large personal and societal impact is further compounded by their co-occurrence, which is associated with more severe symptoms, greater treatment resistance and poorer prognoses (Bair et al., 2003; Roughan et al., 2021). Consequently, recognising and understanding this comorbidity better is critical for improved treatment for those affected.

Over the past two decades, preclinical studies have provided evidence of transdiagnostic molecular mechanisms that help explain the co-occurrence of pain and mood disorders, such as altered serotonin signalling, dysregulated inhibitory-excitatory neurotransmission, and imbalances in inflammation-related cytokines (Humo et al., 2019; Li et al., 2025). Recent neuroimaging studies in humans have also reported converging alterations in brain structure, further supporting the idea that chronic pain and depression share a common neurobiological basis. While there is still little research examining the comorbidity directly, studies of each disorder separately have implicated overlapping changes in key regions across both conditions, including the anterior cingulate cortex (ACC), caudate, and insula (Bhatt et al., 2024; Nakamura et al., 2025; Schmaal et al., 2017; Tagliaferri et al., 2022). These regions are thought to mediate some of the affective, sensory and cognitive aspects of chronic pain and depression. In addition to disorder-specific evidence, a recent meta-analysis of cortical thickness across chronic pain, depression, and anxiety used a conjunctional framework to identify four shared clusters of cortical thinning: the right insula, left ACC, left inferior frontal gyrus (pars triangularis), and left middle temporal gyrus (Yu et al., 2025).

While these common abnormalities in brain structure may reflect genuine transdiagnostic alterations, they may also arise from comorbidity-related confounding in the samples studied (Fortea et al., 2021). Many previous neuroimaging studies of chronic pain and depression, especially those used to infer shared structural differences, have used case-control designs that do not account for pain-depression comorbidity. Given the high co-occurrence of depression and chronic pain, findings from these studies are likely to capture signal from the comorbid subgroup. As a result, when overlapping associations are interpreted as evidence of “shared” brain-structure alterations, it is unclear whether they reflect genuinely overlapping neurobiology or artefacts of comorbidity confounding. Characterising the pattern of structural alterations in the comorbid group, and how this differs from individuals with chronic pain only or depression only, could provide important insights into the pathophysiology of comorbidity and help identify potential biomarkers.

Leveraging neuroimaging data from the UK Biobank (UKB), a large prospective cohort of approximately 500,000 UK participants with extensive health-related data, and its recent milestone of 100,000 participants attending an imaging assessment (Littlejohns et al., 2020), we can directly examine brain structure in individuals with current comorbid chronic pain and depression. Crucially, the scale of this dataset will allow us to stratify participants into comorbidity-sensitive groups, namely those with both chronic pain and depression, those with chronic pain only, those with depression only, and those with neither condition, and to compare all groups while retaining sufficient statistical power to detect small effects.

Therefore, using UKB data, this cross-sectional study aims to characterise brain structure alterations in individuals with comorbid chronic pain, as well as in disorder-specific groups (chronic pain-only and depression-only). To do this, we used pairwise association analysis to establish profiles of brain structure alterations in each disorder group compared to controls, and to identify any group-specific brain structure abnormalities.

## Methods

### Participants

UKB is a large prospective cohort study comprising 501,963 volunteers recruited from the general UK population between 2006 and 2010 (Sudlow et al., 2015). At recruitment, participants completed extensive health and lifestyle assessments, with linkage and follow-up data collection subsequently providing additional phenotyping, including measures relevant to chronic pain and depression, as well as multimodal brain imaging. For the present analyses, we included 71,214 participants with currently available structural brain imaging from the first imaging assessment and complete data on covariates and the current chronic pain and depression phenotypes used in this study. Descriptive statistics of the sample are reported in Table 1.

**Table 1.** Baseline characteristics of participants in the imaging UK Biobank sample by chronic pain and depression status.

| Characteristic | Controls<br>n = 43,926<br>61.7% | Comorbidity<br>n = 1,377<br>1.9% | Chronic pain-only<br>n = 24,981<br>35.1% | Depression-only<br>n = 930<br>1.3% |
| --- | --- | --- | --- | --- |
| Age, years, mean (SD) | 65.9 (7.8) | 63.4 (7.9) | 65.9 (7.9) | 63.1 (7.9) |
| Sex |  |  |  |  |
| • Female | 22,266 (50.7%) | 893 (64.9%) | 14,152 (56.7%) | 529 (56.9%) |
| • Male | 21,660 (49.3%) | 484 (35.1%) | 10,829 (43.3%) | 401 (43.1%) |
| Ethnic background |  |  |  |  |
| • Non-White | 1,344 (3.1%) | 88 (6.4%) | 803 (3.2%) | 64 (6.9%) |
| • White | 42,582 (96.9%) | 1,289 (93.6%) | 24,178 (96.8%) | 866 (93.1%) |
| Townsend deprivation index, mean (SD) | -2.0 (2.7) | -1.0 (3.2) | -1.8 (2.8) | -1.3 (3.0) |
| BMI, kg/m<sup>2</sup>, mean (SD) | 26.0 (4.1) | 28.7 (5.7) | 27.2 (4.8) | 26.7 (4.6) |
| Smoking status |  |  |  |  |
| • Never smoker | 28,463 (64.8%) | 791 (57.4%) | 15,081 (60.4%) | 577 (62.0%) |
| • Current/previous smoker | 15,463 (35.2%) | 586 (42.6%) | 9,900 (39.6%) | 353 (38.0%) |
| Alcohol intake frequency |  |  |  |  |
| • Monthly / never | 11,936 (27.2%) | 657 (47.7%) | 7,965 (31.9%) | 344 (37.0%) |
| • Weekly | 31,990 (72.8%) | 720 (52.3%) | 17,016 (68.1%) | 586 (63.0%) |

BMI = body mass index, SD = standard deviation.

### Current depression phenotype

Current depression was measured using the Patient Health Questionnaire (PHQ)-2 (data field IDs: 2050, 2060) (Abdelhack et al., 2023; See et al., 2025). In UKB, the PHQ-2 items were collected via the UKB Touchscreen Questionnaire at each assessment visit. The PHQ-2 asks participants how often, over the past two weeks, they have been bothered by: (1) little interest or pleasure in doing things, and (2) feeling down, depressed, or hopeless. Responses are recorded on a 4-point Likert scale: 0 = not at all, 1 = several days, 2 = more than half the days, and 3 = nearly every day, yielding a total score range of 0–6. Following established scoring guidelines, participants with PHQ-2 scores ≥ 3 were classified as current depression cases, and those scoring ≤ 2 were classified as controls (Kroenke et al., 2009). The PHQ-2 was used to determine current depression status as it was administered on the same day as the brain scan. Furthermore, in validation work against an independent structured clinical interview, a PHQ-2 cut-off of ≥ 3 demonstrated high sensitivity and specificity (83% and 92%, respectively) for major depression (Kroenke et al., 2003)

### Current chronic pain phenotype

Chronic pain phenotyping was also derived from the UKB Touchscreen Questionnaire on the day of scanning. Participants were asked if pain in a particular body site, or all-over pain, interfered with their usual activity in the past month (data field ID: 6159). Participants could choose multiple individual body sites unless all-over pain was selected, and vice versa. For participants reporting pain, they were then asked if this pain had been present for longer than three months (data field IDs: 3799, 4067, 3404, 3571, 3741, 3414, 3773, 2956). Participants reporting at least one site of chronic pain or widespread chronic pain were classified as having chronic pain, while those reporting no chronic pain were classified as controls.

### Comorbid chronic pain and depression phenotypes

Chronic pain and depression status were combined to create a four-level nominal comorbidity variable at the imaging assessment: (1) comorbidity (current chronic pain and current depression), (2) chronic pain-only (current chronic pain without current depression), (3) depression-only (current depression without current chronic pain) and (4) controls (no current depression and no current chronic pain).

### Brain Imaging

For the analysis carried out in this study, imaging-derived phenotypes (IDPs) that had previously been generated by UKB were used. All imaging data were obtained using a standard Siemens Skyra 3T scanner. More information on imaging protocols followed is available from UKB: https://biobank.ctsu.ox.ac.uk/crystal/crystal/docs/brain_mri.pdf. Grey matter morphometric data consisted of cortical thickness, surface area, and volume measures for 33 bilateral individual regions, as defined by the Desikan-Killiany atlas (Desikan et al., 2006). Subcortical volumes were derived using FIRST (FMRIB’s Integrated Registration and Segmentation Tool), resulting in seven bilateral subcortical region volumes (Patenaude et al., 2011). Diffusion tensor imaging (DTI) tracts were derived using AutoPtx, which calculated fractional anisotropy (FA) and mean diffusivity (MD) in 12 bilateral and three unilateral white matter tracts (de Groot et al., 2013). FA indexes the directional coherence of water diffusion in white matter tracts and MD reflects the overall magnitude of water diffusion (Beaulieu, 2002); lower FA and higher MD are commonly interpreted as indicating lower white matter microstructural integrity (Alexander et al., 2007). All brain measures were scaled to facilitate comparability across metrics. Given known hemispheric asymmetries, left and right hemisphere measures were treated as separate measures and modelled separately. Values more than three standard deviations (SD) from the mean were excluded; counts of removed outliers are provided in Supplementary Table 4.

### Statistical models and covariates

All analyses were conducted in R (v4.4.1) and were restricted to complete cases. Group differences across comorbidity groups (comorbidity, chronic pain-only, depression-only and controls) were assessed using an analysis of covariance (ANCOVA) framework implemented via ordinary least-squares linear regression (lm()), with comorbidity group as the predictor and each brain-structure metric as the outcome. An omnibus F-test was used to evaluate the overall effect of the comorbidity group. Where a significant group effect was observed (p < 0.05), post hoc pairwise comparisons were performed using covariate-adjusted estimated marginal means (emmeans), with false discovery rate (FDR) correction applied across all pairwise comparisons. Statistical significance was predefined as p_FDR_ < 0.05.

All statistical analyses adjusted for age, sex, body mass index (BMI), ethnic background, socioeconomic status, smoking status, alcohol consumption, imaging centre, and three head position coordinates in the scanner. Two age terms were included: age at the time of the initial imaging assessment (data field 21003) and age squared, to capture both linear and non-linear associations between age and brain structure. Given the high proportion of White British participants in UKB, self-reported ethnic background (data field 21000) was coded as a binary variable (White vs non-White). Socioeconomic status was indexed using the Townsend deprivation score (data field 22189). In analyses of cortical structure and subcortical volumes, intracranial volume (ICV; data field 26521) was additionally included to account for overall head size. Full details on covariate sources and coding are provided in Supplementary Tables 1–2.

## Results

### Characteristics of the study population

A flowchart of the study sample is shown in Figure 1. After excluding participants with missing brain structure data, covariates, and/or pain or depression phenotyping, 71,214 participants remained. Of these, 43,926 had neither chronic pain nor depression, and 1,377 had comorbid chronic pain and depression, 24,981 had chronic pain only, and 930 had depression only. Descriptive statistics across key variables are reported in Table 1. Compared with participants in the other groups, those with comorbid chronic pain and depression were more likely to be female (64.9%), have higher deprivation and BMI (28.7 kg/m²), be current or former smokers (42.6%), and were less likely to report weekly alcohol consumption (52.3%). Participants in the depression-only group tended to be younger (mean age 63.1 years) and were more likely to be non-White (6.9%) than participants in the other groups; however, these characteristics were very similar to those observed in the comorbidity group.

**Figure 1.**
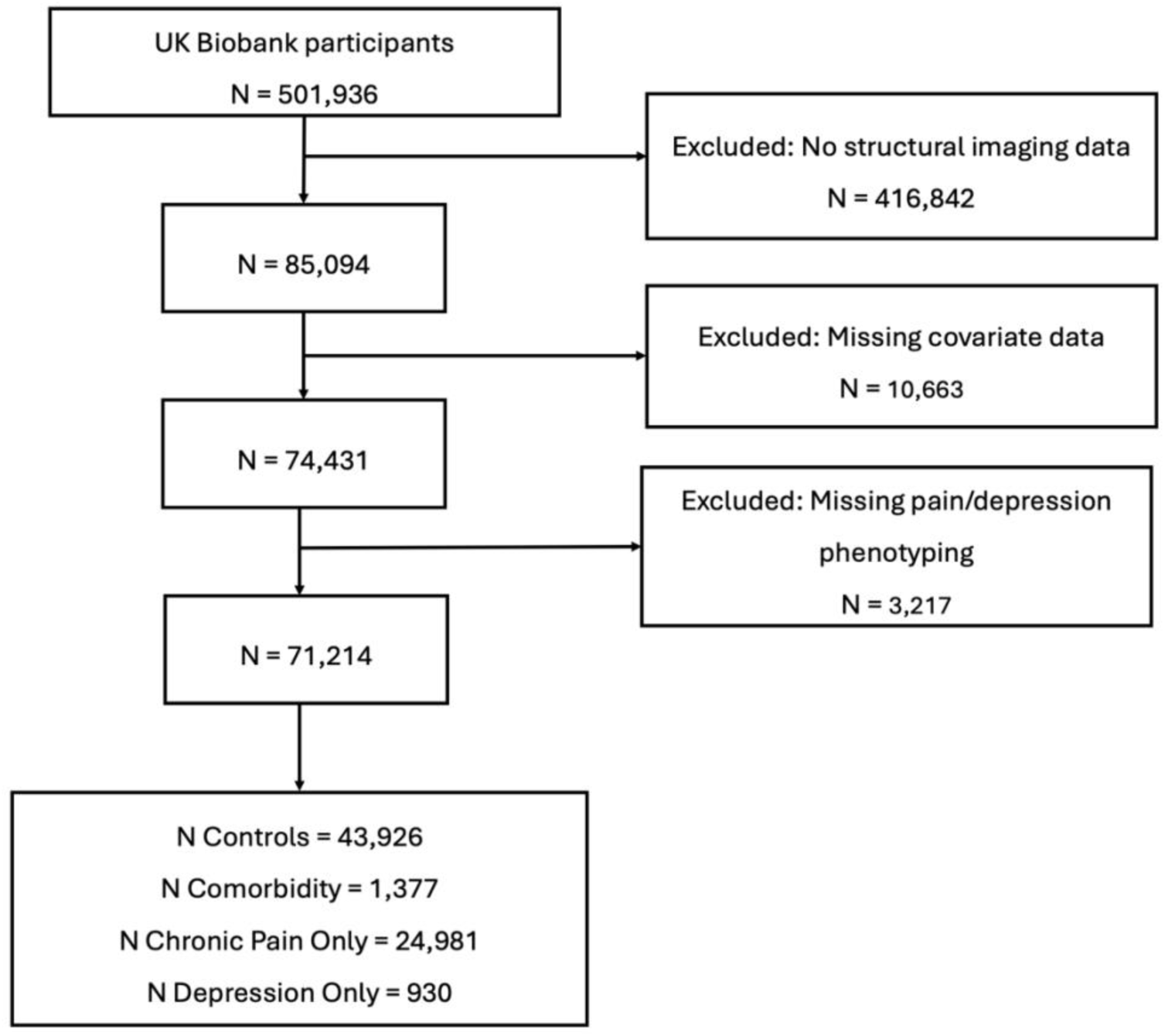
Flow diagram of the study sample.

### Group differences in cortical structure

Pairwise associations between groups were examined for all brain structure measures showing a significant overall group effect; full results are reported in Supplementary Tables 5–6. For clarity, Figure 2 presents the standardised effects of each disorder group (comorbidity, chronic pain-only and depression-only) compared to controls, i.e., those with neither condition.

**Figure 2.**
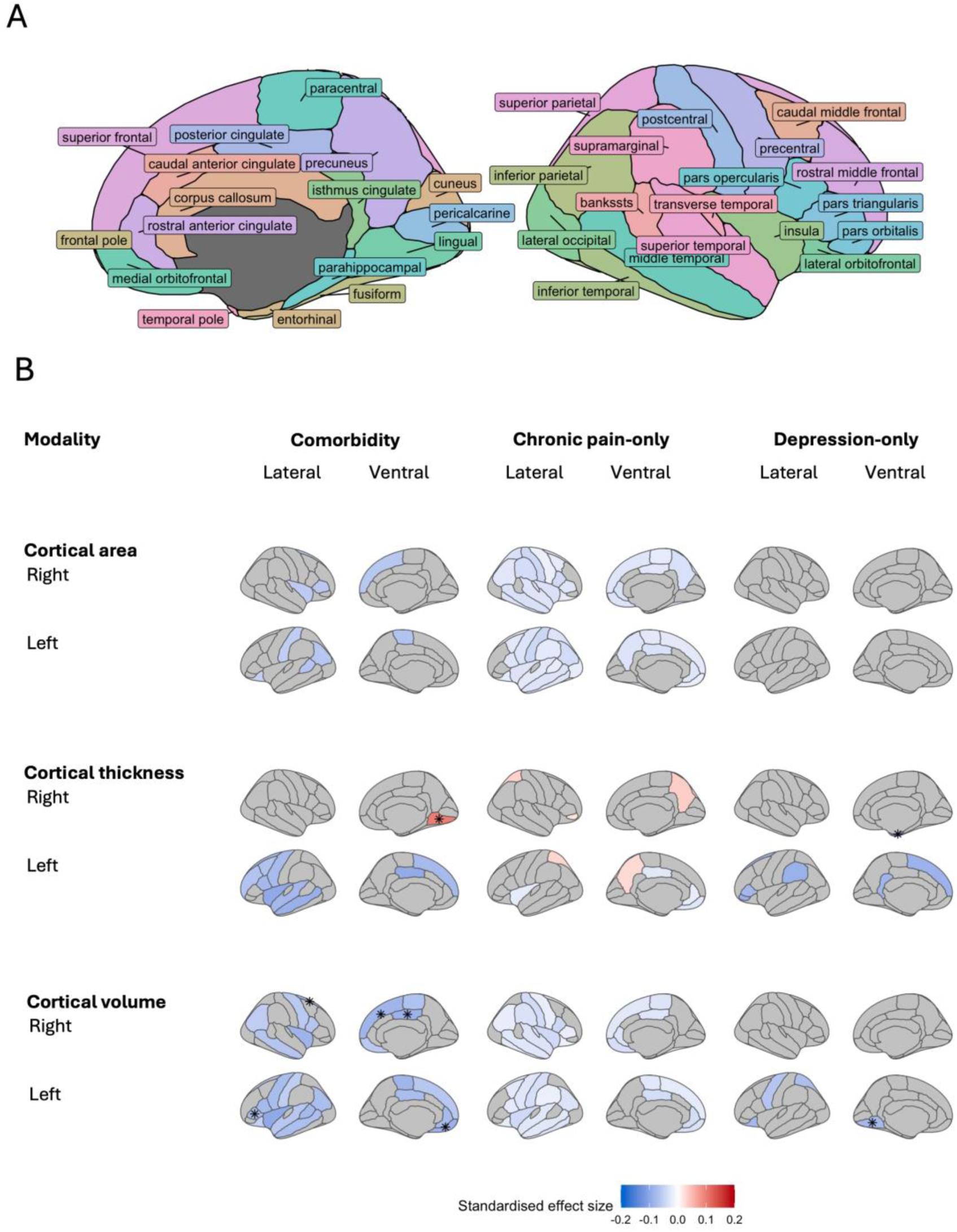
The effect of chronic pain and depression groups on cortical structure. **A** Cortical regions of interest in medial and lateral views. **B** Significant differences in cortical surface area, thickness, and volume for the comorbidity, chronic pain-only and depression-only groups relative to controls. Effect sizes are shown for regions exhibiting significant differences; positive values indicate higher brain metric values relative to controls and negative values indicate lower values (red and blue, respectively). Asterisks (*) denote regions that also differed significantly for the indicated group compared with all other groups.

Compared with controls, the comorbidity group showed lower cortical surface area in the left banks of superior temporal sulcus (bankssts), inferior parietal, lateral orbitofrontal, paracentral, and postcentral cortex, as well as the right insula, pars triangularis, and superior frontal gyrus (β range = −0.065 to −0.049, p_FDR_ < 0.05). Lower cortical thickness was also observed in multiple left-hemisphere frontal and temporal regions (β range = −0.096 to −0.065, p_FDR_ < 0.05). Cortical thickness was greater in the lingual gyrus (β = 0.098, p_FDR_ = 0.002). Widespread lower cortical volume was observed across a broad set of regions (β range = −0.096 to −0.050, p_FDR_ < 0.05). Effects were largely bilateral in frontal regions, including the medial orbitofrontal, paracentral, pars opercularis, precentral, and superior frontal cortices, with additional left-hemisphere effects in the frontal pole, lateral orbitofrontal cortex, and pars triangularis. Lower volume was also observed in parietal regions (bilateral inferior parietal and left postcentral), temporal regions (left bankssts, bilateral middle temporal, and left superior temporal), limbic regions (bilateral posterior cingulate cortex (PCC) and left rostral anterior cingulate), and the insula bilaterally.

Compared with controls, the chronic pain-only group showed widespread lower cortical surface area, with significantly lower surface area observed in 42 of 66 hemisphere-specific regional measures (β range = −0.042 to −0.016, p_FDR_ < 0.05). The largest effects were in the postcentral gyrus (left: β = −0.043, p_FDR_ < 0.001; right: β = −0.042, p_FDR_ < 0.001). More local differences in cortical thickness were observed as greater thickness in bilateral measures of the precuneus and superior parietal lobule, and in the right pars orbitalis (β range = 0.021 to 0.036, p_FDR_ < 0.05). Lower thickness was also observed in the left medial orbitofrontal cortex, PCC, insula, and rostral anterior cingulate cortex (β range = −0.026 to −0.019, p_FDR_ < 0.05). Significantly lower cortical volume was observed in 36 of 66 regions (β range = −0.041 to −0.015, p_FDR_ < 0.05); the largest effects were in the right pars opercularis (β = −0.041, p_FDR_ < 0.001) and bilateral insula (left: β = −0.040, p_FDR_ < 0.001; right: β = −0.040, p_FDR_ < 0.001).

We found no evidence of significant differences in cortical surface area in the depression-only group versus controls. Cortical thickness was significantly lower in the right entorhinal cortex (β = −0.140, p_FDR_ < 0.001) and in several left-hemisphere regions: the pars orbitalis, supramarginal gyrus, superior frontal gyrus, isthmus cingulate cortex, and pars triangularis (β range = −0.091 to −0.079, p_FDR_ < 0.05). Significantly lower cortical volume was also observed in the depression-only group in the left lateral orbitofrontal cortex, lingual gyrus, superior parietal lobule, and precentral gyrus (β range = −0.079 to −0.062, p_FDR_ < 0.05).

In addition to comparing each disorder group with controls, the full set of pairwise contrasts was used to identify structural differences specific to each group. When the comorbidity group was compared with the other groups, right lingual gyrus thickness was greater (β range = 0.086 to 0.105, p_FDR_ < 0.05), and cortical volume was lower in the left pars triangularis (β range = −0.106 to −0.063, p_FDR_ < 0.05), left medial orbitofrontal cortex (β range = −0.086 to −0.054, p_FDR_ < 0.05), right superior frontal cortex (β range = −0.086 to −0.058, p_FDR_ < 0.05) and right PCC (β range = −0.118 to −0.059, p_FDR_ < 0.05). In the depression-only group, cortical thickness was lower in the right entorhinal cortex (β range = −0.142 to −0.107, p_FDR_ < 0.05) and cortical volume was lower in the left lingual gyrus (β range = −0.121 to −0.077, p_FDR_ < 0.05) relative to all other groups.

### Group differences in subcortical volume

Figure 3 shows the standardised effects of each disorder group on subcortical volumes relative to controls. Lower thalamic volume was observed in both the comorbidity group (left: β = −0.048, p_FDR_ = 0.038; right: β = −0.060, p_FDR_ = 0.007) and the depression-only group (left: β = −0.067, p_FDR_ = 0.016; right: β = −0.066, p_FDR_ = 0.015). The comorbidity group also showed lower hippocampal (left: β = −0.062, p_FDR_ = 0.035; right: β = −0.088, p_FDR_ = 0.002) and left accumbens volume (β = −0.073, p_FDR_ = 0.011). There was no evidence of significant subcortical volume differences in the chronic pain-only group compared with the control group.

**Figure 3.**
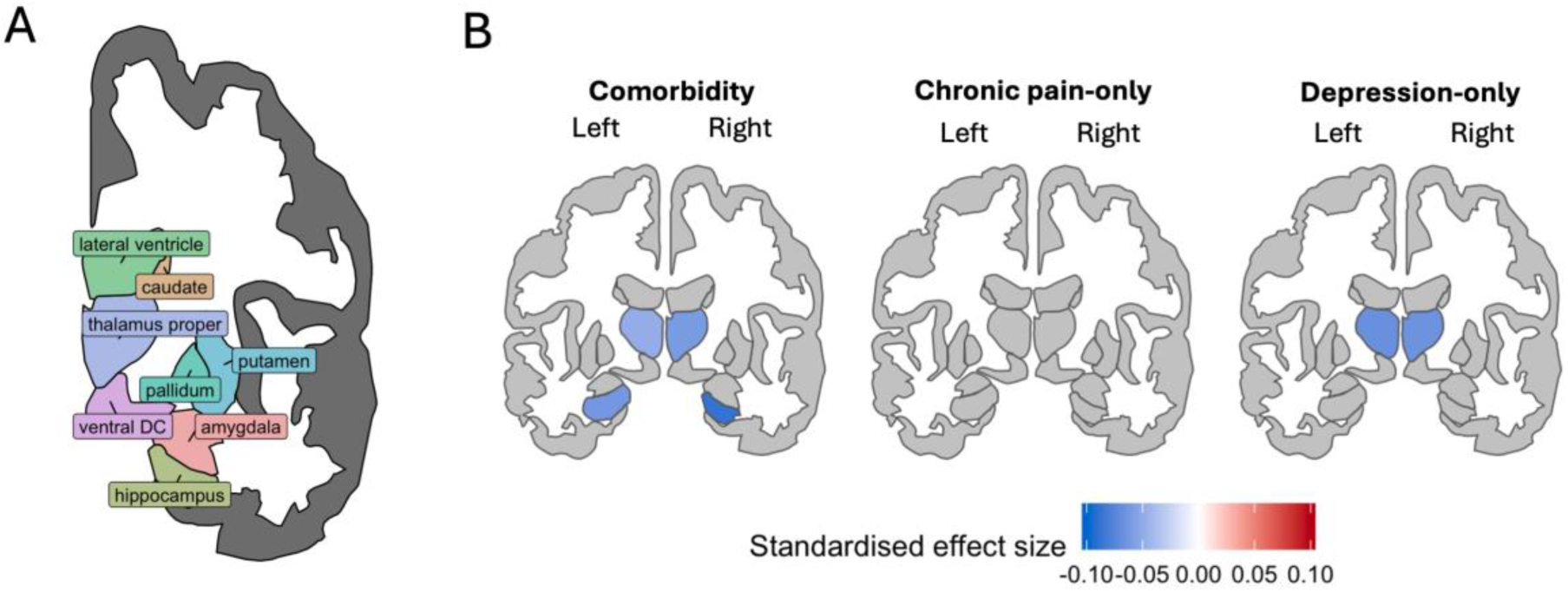
The effect of chronic pain and depression groups on subcortical volume. **A** Subcortical regions of interest in the coronal view. **B** Significant differences in subcortical volume for the comorbidity, chronic pain-only and depression-only groups relative to controls. Effect sizes are shown for regions exhibiting significant differences; negative values indicate lower volumes in each disorder group compared with controls (blue).

### Group differences in measures of white matter microstructure

Figure 4 shows significant group differences in the microstructure of white matter tracts, indexed by FA and MD, compared to controls. The comorbidity group also showed widespread lower FA (15/27 tracts) and higher MD (18/27 tracts), when compared to controls. The largest FA effects were observed in the inferior longitudinal fasciculus (left: β = −0.104, p_FDR_ = 0.001; right: β = −0.116, p_FDR_ < 0.001) and the right posterior thalamic radiation (β = −0.099, p_FDR_ = 0.002). The largest MD effects were observed in the uncinate fasciculus (left: β = 0.122, p_FDR_ < 0.001; right: β = 0.137, p_FDR_ < 0.001) and the right superior thalamic radiation (β = 0.107, p_FDR_ < 0.001). The depression-only group was shown to have significantly lower FA (10/27) and higher MD (15/27) in multiple white matter tracts. The largest FA effects were observed in the left posterior thalamic radiation (β = −0.104, p_FDR_ = 0.007) and the anterior thalamic radiation (left: β = −0.098, p_FDR_ = 0.012; right: β = −0.102, p_FDR_ = 0.010). Largest MD effects were observed in the right corticospinal tract (β = 0.149, p_FDR_ < 0.001) and the left posterior thalamic radiation (β = 0.104, p_FDR_ = 0.001). In the chronic pain-only group, the only significant association was higher FA in the right acoustic radiation (β = 0.022, p_FDR_ = 0.025).

**Figure 4.**
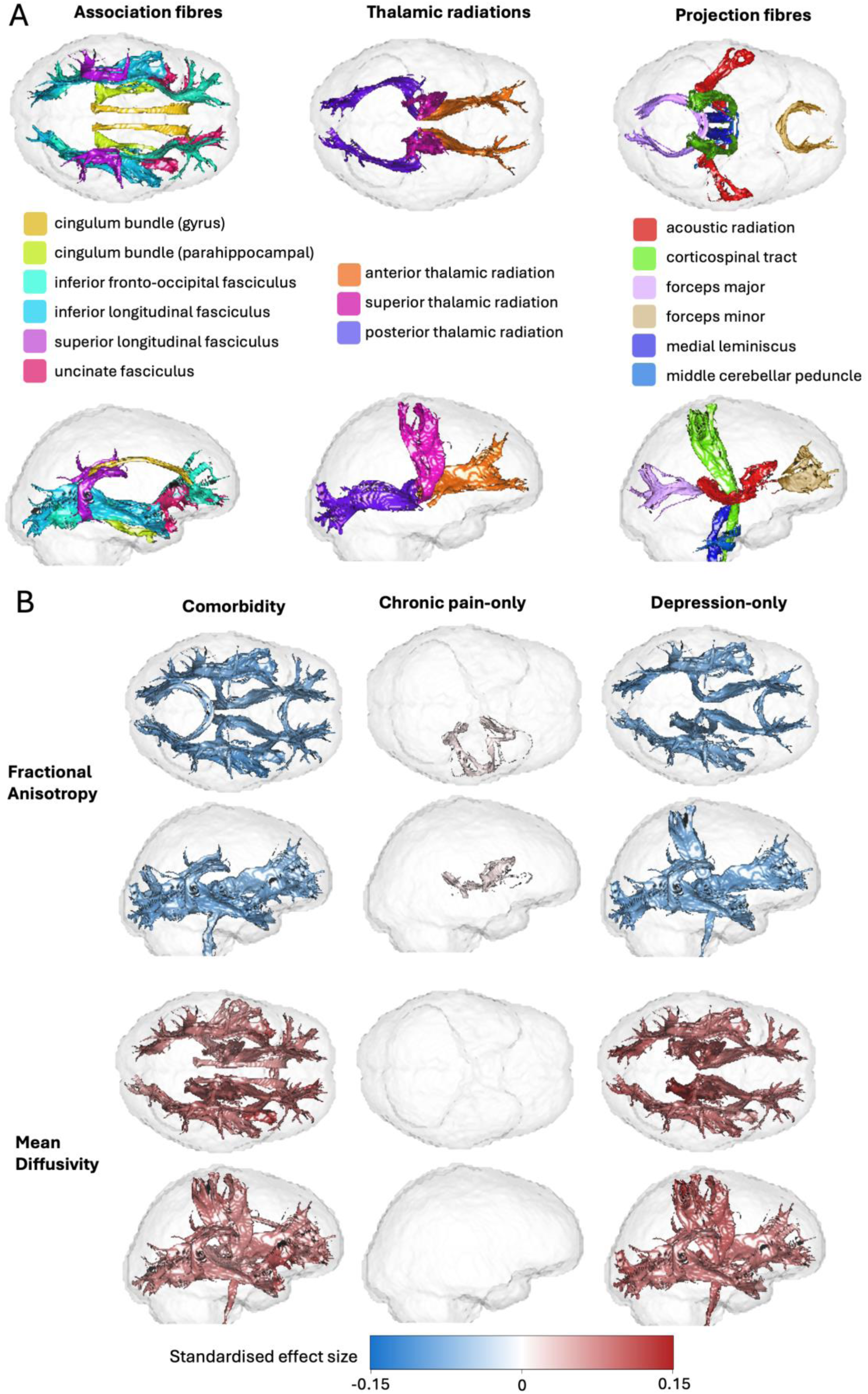
The effect of chronic pain and depression groups on white matter microstructure. **A** White matter tracts of interest in superior and lateral views. **B** Significant differences in measures of white matter microstructure, fractional anisotropy and mean diffusivity for the comorbidity, chronic pain-only and depression-only groups relative to controls. Effect sizes are shown for regions exhibiting significant differences; positive values indicate higher brain metric values relative to controls and negative values indicate lower values (red and blue, respectively).

## Discussion

We examined associations between chronic pain and depression, both as comorbid conditions and as separate disorders, and structural neuroimaging measures in 71,214 UKB participants. Compared with controls, individuals with both chronic pain and depression had local alterations in cortical area and thickness, with more widespread lower cortical volume. Additionally, subcortical volumes of the thalamus, hippocampus and left accumbens volumes were lower, alongside lower FA and higher MD across many white matter tracts, patterns consistent with widespread alterations in white matter microstructure. Relative to controls, the chronic pain-only group had widespread lower cortical surface area and cortical volume, alongside higher FA in the right acoustic radiation. The depression-only group had more circumscribed cortical alterations, limited to several regions with lower cortical thickness and volume. Additionally, thalamic volume was significantly lower, and there was evidence of widespread deficits in white matter microstructure. We also identified brain structures that differed significantly in the comorbidity group compared with all other groups.

A limited number of studies have directly examined brain structure in individuals with comorbid chronic pain and depression. Our findings converge with previous findings from this literature, showing lower volumes in the thalamus, posterior cingulate cortex (PCC), and prefrontal regions (Gustin et al., 2013; Ma et al., 2022). However, there are also inconsistencies, particularly in the hippocampus and anterior insula, where prior work has reported increased volume (Gustin et al., 2013; Ma et al., 2022). The reasons for this discrepancy remain unclear, but one plausible explanation is heterogeneity in pain phenotypes across studies. For example, Gustin *et al*. examined facial pain, and there is evidence that different chronic pain locations are associated with distinct structural brain patterns (Bhatt et al., 2024). It is also important to note that our chronic pain cases included both prolonged pain in discrete body sites and widespread throughout the body. However, previous work has identified differences in symptoms, pain persistence and comorbidities between widespread versus local chronic pain (Viniol et al., 2013), meaning future work separating these phenotypes may be warranted.

Our findings also extend prior cortical analyses by separating cortical surface area and cortical thickness, biologically distinct components of cortical volume (Panizzon et al., 2009), thereby providing a more comprehensive profile of cortical alterations associated with comorbid chronic pain and depression. Using this approach, we identified cortical changes that have not previously been characterised in the comorbidity, including lower surface area in the left postcentral gyrus (primary somatosensory cortex), alongside additional regional area reductions, and lower cortical thickness across left frontal and temporal regions. An additional finding was greater lingual gyrus thickness, which was unique to the comorbid group. The lingual gyrus is part of the visual association cortex, and while its role in pain and depression remains unclear, there is growing evidence that visual cortical regions contribute to pain processing and modulation, potentially via interactions with thalamic and limbic systems (Bush et al., 2021; Wei et al., 2019).

Along with widespread lower cortical volume compared with controls, the comorbid group also showed significantly lower volumes than all other groups in the left pars triangularis, left medial orbitofrontal cortex, right superior frontal cortex, and right PCC. These regions have been implicated in processes relevant to pain-depression comorbidity, including value-based modulation of pain and reward-related pain inhibition (medial orbitofrontal cortex) (Becker et al., 2017), pain catastrophizing and self-referential processing (PCC) (Lee et al., 2018; Zhou et al., 2020) and top-down pain modulation and regulation of negative affect (PFC) (Hiser & Koenigs, 2018; Ong et al., 2019). These unique differences in the comorbidity group suggest that the underlying pathophysiology is driven by, or leads to the development of distinct neural mechanisms, not present in pain or depression as separate disorders. Overall, this indicates a distinct neurobiological profile in individuals with both chronic pain and depression.

Our analysis also extended to DTI-derived FA and MD, which are commonly used as proxy indices of white matter microstructure integrity. To our knowledge, white matter microstructure has not previously been characterised in comorbid chronic pain and depression. Therefore, our results provide a novel insight into widespread structural deficits in many tracts. Notably, two-thirds (66.7%) of the tracts showing significant differences in the comorbidity group also differed in the depression-only group, suggesting both overlapping and distinct white matter microstructural profiles. Much of our current understanding of the neural substrates of comorbid chronic pain and depression comes from preclinical rodent models of persistent pain and accompanying depressive-like behaviours (Li et al., 2025). These studies have delineated specific circuits that mediate the affective and motivational consequences of ongoing pain. However, the translational gap between animal models and human patients and the relative scarcity of well-powered human neuroimaging studies that test and replicate these circuits have limited our understanding of their relevance. By characterising white matter tract microstructure in a large human sample, our work provides a robust and novel perspective on the structural connectivity of pathways implicated in chronic pain and depression comorbidity. Specifically, our findings may reflect structural substrates through which altered coupling between brain regions could contribute to comorbid pain and depression, including fronto-limbic pathways linking ventromedial/orbitofrontal prefrontal regions with the amygdala (uncinate fasciculus) (Liu et al., 2020), thalamo–prefrontal/cingulate pathways (anterior thalamic radiation), and medial network connectivity linking medial PFC/ACC with PCC and other midline regions (cingulum: cingulate gyrus part) (Kummer et al., 2020).

Comparisons between the disorder-specific groups and the comorbidity group allow us to examine how brain structure in the comorbidity group differs from profiles associated with either condition alone, and to identify comorbidity-specific alterations (as discussed above). Contrasts between each disorder-specific group and healthy controls also provided well-powered estimates of disorder-specific differences. Relative to healthy controls, individuals with chronic pain only showed widespread reductions in cortical surface area and cortical volume, consistent with previous UKB analyses in chronic pain based on an earlier imaging release (Bhatt et al., 2024). While white matter microstructure has been examined across chronic pain conditions, the literature remains heterogeneous and is often limited by modest sample sizes and methodological variability (Bautin et al., 2025). This study, therefore, extends current evidence by providing a large-scale characterisation of white matter microstructure differences associated with chronic pain. The only significant DTI result was higher FA in the right acoustic radiation. The acoustic radiation is a thalamocortical pathway that conveys auditory information from the medial geniculate nucleus to primary auditory cortex (Siegbahn et al., 2022). This finding may be relevant to sound hypersensitivity reported in some chronic pain conditions, including fibromyalgia and CRPS-related dystonia (de Klaver et al., 2007; Staud et al., 2021), and to phonophobia, which is a common feature of migraine (Headache Classification Committee of the International Headache Society, 2018).

The cortical and subcortical differences observed in the current depression-only group partly diverged from patterns reported in previous UKB studies of depression (Harris et al., 2022; Johns et al., 2025; Shen et al., 2017). These discrepancies may reflect differences in phenotyping. For example, Harris et al. examined the effect of three lifetime depression phenotypes on brain structure in UKB, whereas our depression definition reflects current symptoms, which may be more sensitive to state-related or transient changes (Harris et al., 2022). These discrepancies highlight lower left lingual cortex volume, left supramarginal gyrus volume, and thalamus volume as potential transient changes associated with current depressive state, although further work is needed to verify this. Notably, the pattern of cortical thinning observed in the left hemisphere is consistent with a smaller case-control study (n = 61) reporting reduced thickness in left pars triangularis, pars orbitalis, and supramarginal gyrus in participants currently experiencing a depressive episode (Wang et al., 2024). Despite using a different depression phenotype, we observed disrupted white matter microstructural measures, broadly consistent with prior UKB reports showing widespread white matter microstructure alterations in depression (Harris et al., 2022; Shen et al., 2017). These findings are consistent with conceptualisations of depression as a disorder of network-level functional dysconnectivity and suggest that white-matter microstructural alterations may provide a structural substrate that could contribute to this dysconnectivity (Chai et al., 2023; Gong & He, 2015).

The present findings provide a robust characterisation of brain structural differences associated with comorbid chronic pain and depression; however, the temporal ordering of these associations remains unclear. Leveraging repeat imaging in UKB and other cohorts, longitudinal analyses could test whether these structural differences precede the onset of chronic pain and depression, emerge following symptom development, or change with symptom persistence and severity. In addition, converging evidence from longitudinal and genetic approaches suggests a bidirectional relationship between pain and depression, indicating that each may increase risk for the other (Jiang et al., 2025; Johnston et al., 2019; Zhao et al., 2023). Longitudinal imaging could therefore be used to test whether specific brain structural changes mediate the pathway from chronic pain to depression, and vice versa, helping to identify candidate mechanisms that contribute to the development and maintenance of comorbidity.

Interpretation of the results is limited by our inability to disentangle the effects of symptom severity from the effects of comorbidity on brain structure. On average, individuals with both chronic pain and depression experience greater symptom burden than those with either condition alone (Bair et al., 2003), meaning the effects we observed in the comorbidity group could reflect disorder severity rather than comorbidity-specific effects. Additionally, the study relies on self-reported, broad definitions of chronic pain and depression, which may introduce bias, increase variability, and limit the portability of the findings. A further limitation is that the cross-sectional design precludes inferences about causality or temporal ordering. In addition, because pain is a symptom of many neurological conditions, participants with neurological disorders were not excluded. This likely increases generalisability, but it may also introduce heterogeneity and potential confounding that should be considered when interpreting the findings. Similarly, other potential comorbidities, such as diabetes and arthritis, and medication status were not controlled for, which may associate with an individual’s risk of both chronic pain and depression, and brain structure, biasing results. Finally, UKB is subject to a “healthy volunteer” selection bias that is even more pronounced in the imaging subset and may limit representativeness and attenuate effect sizes if individuals with more severe illness are underrepresented (Fry et al., 2017; Keyes & Westreich, 2019; Lyall et al., 2022).

## Conclusion

Our study characterises brain structure differences in comorbid chronic pain and depression, alongside disorder-specific profiles for chronic pain-only and depression-only groups, in the largest single neuroimaging sample to date. Comorbid chronic pain and depression showed a distinct pattern of brain structural alterations characterised by regional differences in cortical surface area and thickness, widespread reductions in cortical volume, lower thalamic, hippocampal and left accumbens volumes, and widespread alterations in measures of white matter microstructure. In addition, comparisons with the disorder-specific groups identified alterations unique to the comorbidity group, supporting a distinct neuroanatomical profile. Disorder-specific analyses showed widespread reductions in cortical surface area and volume in the chronic pain-only group, and region-specific cortical and subcortical differences alongside evidence of widespread deficits in white matter microstructure in the depression-only group. Overall, these findings advance understanding of the neurobiological correlates of comorbid chronic pain and depression and may have an important role in clinical management, not only for application in developing more targeted interventions but also in patient information and education.

## Data availability

Data from UK Biobank (application number 4844) were used in this study. Researchers can apply to use UK Biobank data for health-related research that is in the public interest. Further information is available from the [UK Biobank website](https://www.ukbiobank.ac.uk/register-apply/).

## Code availability

The code used to perform analyses for this study will be made publicly available after publication at https://github.com/hannahcasey/UKB_brain_structure_chronic_pain_depression.

## Ethics and consent

The UK Biobank study was approved by the North West Multi-centre Research Ethics Committee (REC reference: 11/NW/0382). Written informed consent was obtained from each participant.

## Author contributions

**Hannah Casey:** Conceptualisation; data curation; formal analysis; methodology; software; validation; visualisation; writing - original draft; writing - review & editing

**Mark J. Adams:** Data curation, resources

**Andrew M. McIntosh:** Funding acquisition

**Marie T. Fallon:** Conceptualisation; supervision; writing - review & editing

**Daniel J. Smith:** Conceptualisation; supervision; writing - review & editing

**Rona J. Strawbridge:** Conceptualisation; methodology; supervision; writing - review & editing

**Heather C. Whalley:** Conceptualisation; methodology; supervision; writing - review & editing

